# From Symptoms to Strategy: Pre-procedural NYHA Class as a Key to Risk Stratification and Personalized TAVR Management

**DOI:** 10.1101/2025.05.19.25327961

**Authors:** Birgit Markus, Philipp Lauten, Georgios Chatzis, Leonard Steinheisser, Torben Zaun, Nikolaos Patsalis, Styliani Syntila, Harald Lapp, Bernhard Schieffer, Julian Kreutz

**Affiliations:** Philipps University of Marburg, Germany; University Hospital of Marburg, Department of Cardiology, Angiology, and Intensive Care Medicine, Germany; Central Hospital of Bad Berka, Department of Cardiology, Bad Berka, Germany

**Author notes:** Corresponding author: Birgit Markus, MD, Philipps University of Marburg, University Hospital, Department of Cardiology, Angiology, and Intensive Care Medicine, Baldingerstrasse, 35043 Marburg.

**Keywords:** Transcatheter aortic valve replacement (TAVR), New York Heart Association (NYHA), risk stratification, personalized therapy, outcome

## Abstract

**Background:** Transcatheter aortic valve replacement (TAVR) is the standard treatment for aortic stenosis (AS), particularly in elderly or high-risk surgical patients. The New York Heart Association (NYHA) classification, which assesses the severity of heart failure (HF), is a key factor influencing TAVR outcomes. However, its impact on procedural success, complications, and outcomes remains underrepresented in recent studies.

**Methods:** In this multicenter study, data from 2,256 patients who underwent TAVR between 2017 and 2022 at two high-frequency German Heart Centers were analyzed. Demographics, comorbidities, and peri-procedural parameters were evaluated to determine the influence of pre-procedural NYHA classification on complications, hospital stay, and outcomes.

**Results:** Pre-procedural NYHA III/IV were associated with higher peri-procedural complication rates, prolonged hospital stays, and increased mortality compared to NYHA I/II. Specifically, rates were higher for cardiopulmonary resuscitation (5.3% vs. 0.7%; p<0.001), acute coronary intervention (1.9% vs. 0.0%; p=0.006), vasopressor use >6h (11.7% vs. 1.6%; p<0.001), and renal replacement therapy (6.8% vs. 0.2%; p<0.001). Procedure-related complications like vascular closure device failure (4.9% vs. 1.3%; p=0.008), need for vascular surgery (9.0% vs. 6.3%; p=0.002), and blood transfusion (9.4% vs. 4.7%; p=0.017) were more common in NYHA IV. Median hospital stay was longer in NYHA IV (10.0 vs. 6.0 days; p<0.001). The 30-day mortality rate was 8.3% (NYHA IV) vs. 1.4% (NYHA I/II), and 1-year mortality was 19.2% vs. 5.2% (p<0.001).

**Conclusions:** NYHA classification is a reliable predictor of TAVR outcomes, highlighting a distinct gradation in risk and recovery profiles associated with higher NYHA classes. The findings emphasize the critical role of comprehensive pre-procedural evaluation and timely intervention. These insights could shape future guidelines on patient selection and management in TAVR, ultimately enhancing survival rates and quality of life, particularly in high-risk patients.

## 1. Introduction

In recent years, transcatheter aortic valve replacement (TAVR) has emerged as a commonly adopted interventional approach for the treatment of aortic stenosis (AS). Although initially reserved for high-risk or inoperable patients, the procedure has rapidly evolved, driven by increasingly promising data since its introduction into clinical practice.^1^ Today, TAVR is broadly accepted and also utilized in patients with intermediate or low risk.^2,3^ However, with demographic shifts and an aging population, nowadays the majority of TAVR patients are octogenarians or even older. Outcomes in this increasingly heterogeneous patient cohort are shaped by various factors, with pre-procedural clinical status remaining a major predictor.^4–6^

Pathophysiological mechanisms triggered by AS are characterized by considerable complexity, resulting in varying severity of heart failure (HF) and left ventricular systolic dysfunction, which can be routinely assessed and quantified using the *New York Heart Association* (NYHA) classification. The development of HF in the context of untreated AS is a gradual process that typically evolves over several years before becoming clinically apparent. In contrast, the correction of AS via TAVR reduces left ventricular afterload, improves hemodynamics, promotes reverse remodeling, alleviates HF symptoms, and enhances both survival and quality of life.^7–10^ The timing of the TAVR procedure, therefore, appears to be a clinically relevant factor even in well-characterized patient subgroups.^11–14^ Major TAVR trials, including the PARTNER and CoreValve trials, as well as large registry data, such as TVT and FRANCE-2, consistently show significant improvements in patient outcomes in dependence on different NYHA classifications.^2,3,15–18^

Although the association between pre-operative NYHA class and outcomes following cardiac surgery is well-established and plays a crucial role in therapeutic decision-making, its specific impact on TAVR success and early complication rates remains unclear. Notably, this factor has not yet been fully integrated into the decision-making process for TAVR.^19,20,21^ To gain a deeper understanding, it is crucial to thoroughly investigate how pre-procedural cardiac decompensation, as reflected by the NYHA classification, influences outcomes after TAVR. This investigation will be pivotal for optimizing patient selection and periprocedural management in an aging population, ultimately improving both individual patient outcomes and the overall safety and effectiveness of the procedure. Moreover, as the aging population grows, healthcare systems face increasing economic pressures, underscoring the importance of efficient and personalized management strategies. The current study seeks to contribute to closing this knowledge gap in medical practice. By investigating the impact of the symptom-based pre-procedural NYHA classification on peri-interventional complications and outcomes, it aims to provide a more nuanced understanding of the predictive value of the pre-procedural NYHA classification in the context of TAVR.

## 2. Materials and methods

### Study design

This study retrospectively analyzed data from patients undergoing TAVR at two German Heart Centers, the University Hospital of Marburg and the Central Hospital of Bad Berka, between January 2017 and December 2022. Inclusion criteria were TAVR procedure and documented pre-procedural NYHA classification as an indicator of the severity of cardiac decompensation status and HF. Peri-procedural outcomes and data from various domains, including physician reports, laboratory results, admission electrocardiograms (ECGs), imaging studies, and peri-procedural notes, were compared across different NYHA classes. Therefore, patients were categorized based on their pre-TAVR NYHA classification, with classes I and II combined and compared separately to classes III and IV.

### Statistical analysis

Differences between NYHA classes were analyzed with various statistical tests processed by SPSS software (version 29) and GraphPad Prism (version 10.1). Significance was set at p ≤0.05. For categorical variables, Pearson’s chi-squared test was applied unless the expected frequencies were low, in which case Fisher’s exact test was employed. Continuous variables were examined using one-way ANOVA with Welch’s correction for unequal variances or the Kruskal-Wallis test for non-normal distributions. Post-hoc pairwise comparisons were conducted with appropriate adjustments for multiple testing, including the Bonferroni correction to reduce type I errors and the Games-Howell test for unequal variances. The chi-squared n-1 test was applied for adjusted categorical analyses.

### Ethics

The study was approved by the Ethics Committee of Philipps University of Marburg (reference RS 22/65, approved on October 27, 2022) and the Ethics Committee of the Medical Association of Thuringia (approved on July 13, 2023). Both approvals were obtained following the principles of the Declaration of Helsinki.

## 3. Results

Between January 2017 and December 2022, 2,256 patients underwent the TAVR procedure. NYHA classification status on admission was documented in 2,161 patients. The 30-day all-cause mortality rate of the entire NYHA cohort was 3.1%, and the 1-year all-cause mortality rate was 8.9%. Patients with NYHA IV were significantly older, with a mean age of 80.5 ± 6.3 years compared to those with NYHA I/II and III classification (I/II: 79.1 ± 5.9 years, III: 79.7 ± 6.4 years; p=0.013). A comprehensive listing of demographics and comorbidities is provided in Table 1.

**Table 1.** Demographic data and comorbidities are stratified by NYHA classification at admission. Abbreviations: n: number of patients with valid data; BMI: body mass index; CHD: coronary heart disease; PAD: peripheral arterial disease; NYHA: New York Heart Association; py: pack years; KDIGO: Kidney Disease Improve Global Outcomes; COPD: chronic obstructive pulmonary disease. ^1^: n (%); ^2^: Mean (SD).

|  | n | NYHA I/II | NYHA III | NYHA IV | p-value |
| --- | --- | --- | --- | --- | --- |
| Number of patients | 2161 | 558 | 1337 | 266 |  |
| Age (years)<sup>2</sup> | 2161 | 79.1 ($\pm 5.9$) | 79.7 ($\pm 6.4$) | 80.5 ($\pm 6.3$) | **0.013** |
| Male sex<sup>1</sup> | 2161 | 319 (57.2) | 699 (52.3) | 137 (51.5) | 0.120 |
| BMI ($\text{kg}/\text{m}^2$)<sup>2</sup> | 2154 | 27.9 ($\pm 4.9$) | 28.7 ($\pm 5.3$) | 28.6 ($\pm 5.6$) | **0.006** |
| Cardiac bypass surgery<sup>1</sup> | 2161 | 34 (6.1) | 84 (6.3) | 16 (6.0) | 0.978 |
| Atrial fibrillation<sup>1</sup> | 2161 | 158 (28.3) | 502 (37.5) | 121 (45.5) | **<0.001** |
| CHD<sup>1</sup> | 2161 | 290 (52.0) | 761 (56.9) | 173 (65.0) | **0.002** |
| PAD ≥ stage 2<sup>1</sup> | 2160 | 41 (7.3) | 210 (15.7) | 39 (14.7) | **<0.001** |
| Arterial hypertension<sup>1</sup> | 2160 | 486 (87.1) | 1201 (89.9) | 241 (90.6) | 0.151 |
| Hyperlipidemia<sup>1</sup> | 2159 | 390 (70.0) | 909 (68.0) | 180 (67.7) | 0.666 |
| Diabetes mellitus<sup>1</sup> | 2161 | 205 (36.8) | 533 (39.9) | 130 (48.9) | **0.004** |
| Insulin-dependent diabetes mellitus<sup>1</sup> | 2160 | 86 (15.4) | 232 (17.4) | 59 (22.2) | 0.057 |
| Nicotine abuse ($> 5\text{py}$)<sup>1</sup> | 2161 | 81 (14.5) | 218 (16.3) | 40 (15.0) | 0.592 |
| Chronic renal failure<br>KDIGO ≥ stage 3<sup>1</sup> | 2161 | 179 (32.1) | 593 (44.4) | 171 (64.3) | **<0.001** |
| Renal replacement therapy<sup>1</sup> | 2161 | 11 (2.0) | 19 (1.4) | 9 (3.4) | 0.085 |
| COPD ≥ GOLD 2<sup>1</sup> | 2160 | 37 (6.6) | 166 (12.4) | 40 (15.0) | **<0.001** |
| Cerebral stroke<sup>1</sup> | 2158 | 20 (3.6) | 58 (4.3) | 15 (5.6) | 0.401 |
| Malignant disease<sup>1</sup> | 2160 | 33 (5.9) | 142 (10.6) | 34 (12.8) | **0.001** |
| Pacemaker<sup>1</sup> | 2161 | 45 (8.1) | 158 (11.8) | 41 (15.4) | **0.005** |

The subsequent analysis focused on pre-interventional parameters, including ECGs, chest X-rays, laboratory findings, and echocardiography, as summarized in Table 2. Notably, pre-interventional ECGs showed no significant differences between NYHA classes. However, chest X-rays revealed progressively worsening signs of pulmonary venous congestion, pleural effusions, and infiltrates with increasing NYHA stage. Laboratory analyses demonstrated progressively higher levels of C-reactive protein (CRP) and hemoglobin A1c (HbA1c), along with decreased hemoglobin and hematocrit levels, elevated leukocyte counts, and significantly higher N-terminal pro-B-type natriuretic peptide (NT-proBNP) concentrations correlating with advanced NYHA stages. Echocardiography showed a significantly higher prevalence of at least moderately reduced left ventricular ejection fraction (LVEF ≤45%) and advanced concomitant valvular heart disease (grade 2/3 mitral and/or tricuspid regurgitation) in patients with higher NYHA classifications. Moreover, the peak flow velocity across the aortic valve (Vmax) and mean aortic gradient (ΔPmean) were found to decrease progressively with increasing NYHA stage, indicating more low flow and low gradient AS in advanced stages of HF (Table 2).

**Table 2.** Pre-interventional diagnostic examinations in different NYHA classes. Abbreviations: n: number of patients with valid data; bpm: beats per minute; AV Block: atrioventricular block; CRP: C-reactive protein; HbA1c: hemoglobin A1c, NT-proBNP: N-terminal prohormone of brain natriuretic peptide; LVEF: left ventricular ejection fraction; Vmax: peak transvalvular velocity of aortic valve; ΔPmean: mean aortic valve pressure gradient. ^1^: n (%); ^2^: Mean (SD), ^3^: Median [IQR].

|  | n | NYHA I/II | NYHA III | NYHA IV | p-value |
| --- | --- | --- | --- | --- | --- |
| Number of patients | 2161 | 558 | 1337 | 266 |  |
| **Pre-interventional ECG** |  |  |  |  |  |
| Heart rate (bpm)<sup>4</sup> | 2161 | 74.2 ± 14.1 | 75.6 ± 14.9 | 77.5 ± 16.6 | 0.103 |
| Right bundle branch block<sup>1</sup> | 2161 | 53 (9.5) | 97 (7.3) | 14 (5.3) | 0.076 |
| Left bundle branch block<sup>1</sup> | 2161 | 48 (8.6) | 112 (8.4) | 28 (10.5) | 0.522 |
| First degree AV block<sup>1</sup> | 2161 | 62 (11.1) | 126 (9.4) | 25 (9.4) | 0.514 |
| **Pre-interventional chest X-ray** |  |  |  |  |  |
| Pneumonia/ infiltrates | 2161 | 1 (0.2) | 36 (2.7) | 15 (5.6) | **<0.001** |
| Pleural effusion<sup>1</sup> | 2130 | 31 (5.6) | 137 (10.2) | 60 (22.6) | **<0.001** |
| Other congestion signs<sup>1</sup> | 2161 | 19 (3.4) | 87 (6.5) | 27 (10.2) | **<0.001** |
| **Pre-interventional laboratory parameters** |  |  |  |  |  |
| CRP (mg/dL)<sup>3</sup> | 2155 | 2.3 [1.0 – 5.1] | 3.3 [1.4 – 8.7] | 6.2 [2.0 – 15.4] | **<0.001** |
| HbA1c (%)<sup>3</sup> | 1739 | 5.9 (5.5 – 6.4) | 6.0 (5.6 – 6.6) | 6.1 (5.6 – 6.8) | **<0.001** |
| Hemoglobin (g/L)<sup>3</sup> | 2161 | 130 (119 - 140) | 127 (114 - 137) | 122 (109 - 130) | **<0.001** |
| Hematocrit (L/L)<sup>3</sup> | 2158 | 0.39 (0.36 – 0.41) | 0.38 (0.35 – 0.41) | 0.37 (0.34 – 0.40) | **<0.001** |
| Leukocyte count (g/L)<sup>3</sup> | 2151 | 7.5 (6.3 – 8.8) | 7.7 (6.5 – 9.3) | 8.2 (6.6 – 9.9) | **<0.001** |
| Platelet count (g/L)<sup>3</sup> | 2161 | 203 (169 - 245) | 210 (171 - 258) | 209 (164 - 262) | 0.129 |
| Sodium (mmol/L)<sup>3</sup> | 2143 | 141 (139 - 143) | 140 (138 - 143) | 141 (138 - 143) | **0.045** |
| NT proBNP (pg/mL)<sup>3</sup> | 1633 | 968 (482 – 2309) | 1391 (464 – 3332) | 3315 (1397 – 8477) | **<0.001** |
| **Pre-interventional echocardiography** |  |  |  |  |  |
| LVEF ≤45%<sup>1</sup> | 2139 | 93 (16.8) | 317 (24.0) | 99 (37.4) | **<0.001** |
| Vmax aortic valve (mmHg)<sup>2</sup> | 1630 | 4.3 (3.9 – 4.6) | 4.1 (3.8 – 4.5) | 4.0 (3.4 – 4.4) | **<0.001** |
| ΔPmean (mmHg)<sup>2</sup> | 1929 | 47.0 (39.0 – 57.0) | 46.0 (37.0 – 60.0) | 43 (32.0 – 55.0) | **0.005** |
| Aortic Valve Area (cm<sup>2</sup>)<sup>3</sup> | 2024 | 0.75 (±0.17) | 0.75 (±0.19) | 0.73 (±0.19) | 0.295 |
| Concomitant vitium (grade 2/3) of the mitral valve<sup>1</sup> | 2133 | 151 (27.5) | 384 (29.1) | 118 (45.0) | **<0.001** |
| Concomitant vitium (grade 2/3) of the tricuspid valve<sup>1</sup> | 2134 | 94 (17.1) | 270 (20.4) | 70 (26.7) | **0.006** |
| Bicuspid aortic valve<sup>1</sup> | 2139 | 60 (10.8) | 107 (8.1) | 22 (8.3) | 0.162 |

The median of the pre-interventional EuroSCORE II score was 3.8 (IQR 2.4-6.7) in NYHA I/II, 4.5 (IQR 2.8-8.2) in NYHA III, and 7.6 (IQR 4.6-14.7) in NYHA IV (p<0.001). Similarly, the median *Society of Thoracic Surgeons* (STS) score was 4.3 (IQR 2.0-10.0) in NYHA I/II, 4.0 (IQR 2.5-9.3) in NYHA III, and 7.0 (IQR 3.8-12.0) in NYHA IV (p<0.001) (Figure 1).

**Figure 1.**
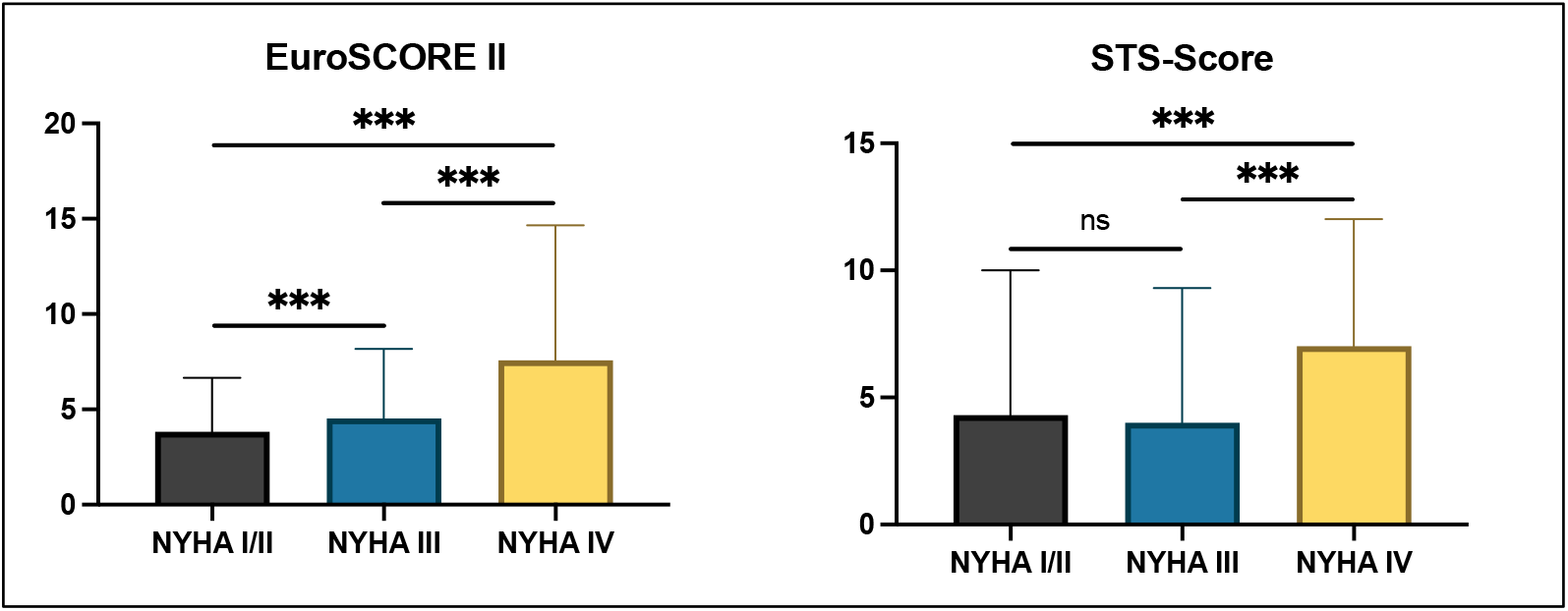
Pre-interventional documentation of EuroSCORE II and STS Score in different NYHA classes. ns: not significant, ***p<0.001.

To evaluate peri-interventional complications associated with TAVR, pre-procedural NYHA classifications were categorized, indicating a significant association between higher pre-procedural NYHA classes and increased peri-interventional complications. In particular, the incidence of peri-interventional cardiopulmonary resuscitation increased with higher NYHA class: 0.7% in NYHA I/II, 3.0% in NYHA III, and 5.3% in NYHA IV (p<0.001). Similarly, the need for acute coronary intervention due to peri-interventional myocardial infarction was also correlated with higher NYHA classes (0.0 % in NYHA I/II, 0.8% in NYHA III, and 1.9% in NYHA IV; p=0.006). In addition, prolonged need for vasopressor therapy (>6h post-procedure) to maintain a mean arterial pressure of about 65 mmHg was more common in patients with higher NYHA classes (1.6% in NYHA I/II, 5.5% in NYHA III, and 11.7% in NYHA IV; p<0.001). The incidence of renal failure requiring temporary renal replacement therapy (RRT) was 0.2% in NYHA I/II, 2.1% in NYHA III, and 6.8% in NYHA IV (p<0.001). Patients classified as NYHA IV experienced the highest rate of vascular closure device failure (4.9%), significantly higher than the 1.3% observed in NYHA I/II and 2.8% in NYHA III (p=0.008). Additionally, NYHA IV patients required the highest frequency of vascular surgery, with rates of 6.3% in NYHA I/II, 4.5% in NYHA III, and 9.0% in NYHA IV (p=0.002). Required blood transfusion increased with advanced NYHA classes (4.7% in NYHA I/II, 7.9% in NYHA III, and 9.4% in NYHA IV; p=0.017) These findings are summarized in Table 3.

**Table 3.** Peri-procedural complications in different NYHA classes. Abbreviations: MI: myocardial infarction; SAVR: surgical aortic valve replacement; RRT: renal replacement therapy.^1^: n (%).

|  | n | NYHA I/II | NYHA III | NYHA IV | p-Wert |
| --- | --- | --- | --- | --- | --- |
| Number of patients | 2161 | 558 | 1337 | 266 |  |
| Cardiopulmonary resuscitation<sup>1</sup> | 2160 | 4 (0.7) | 30 (2.2) | 14 (5.3) | **<0.001** |
| Aortic dissection<sup>1</sup> | 2161 | 1 (0.2) | 5 (0.4) | 0 (0.0) | 0.853 |
| Acute coronary intervention for periprocedural MI<sup>1</sup> | 2161 | 0 (0.0) | 11 (0.8) | 5 (1.9) | **0.006** |
| Switch to SAVR<sup>1</sup> | 2161 | 1 (0.2) | 4 (0.3) | 1 (0.4) | 0.713 |
| Need for vasopressor therapy (> 6 hours post intervention)<sup>1</sup> | 2160 | 9 (1.6) | 74 (5.5) | 31 (11.7) | **<0.001** |
| New onset atrial fibrillation<sup>1</sup> | 2161 | 12 (2.2) | 41 (3.1) | 7 (2.6) | 0.520 |
| Need for pacemaker implantation<sup>1</sup> | 2158 | 70 (12.6) | 204 (15.3) | 38 (14.3) | 0.319 |
| Pericardial tamponade<sup>1</sup> | 2161 | 12 (2.2) | 32 (2.4) | 10 (3.8) | 0.363 |
| Renal failure requiring RRT<sup>1</sup> | 2161 | 1 (0.2) | 28 (2.1) | 18 (6.8) | **<0.001** |
| Dissection of the femoral artery<sup>1</sup> | 2161 | 6 (1.1) | 9 (0.7) | 4 (1.5) | 0.335 |
| Failure of the vascular closure device<sup>1</sup> | 2161 | 7 (1.3) | 37 (2.8) | 13 (4.9) | **0.008** |
| Need for vascular surgery<sup>1</sup> | 2161 | 20 (6.3) | 60 (4.5) | 24 (9.0) | **0.002** |
| Requirement for blood transfusion within the first 48 hours following TAVR<sup>1</sup> | 2161 | 26 (4.7) | 105 (7.9) | 25 (9.4) | **0.017** |
| Cerebral stroke<sup>1</sup> | 2161 | 13 (2.3) | 27 (2.0) | 8 (3.0) | 0.779 |

To evaluate post-procedural outcomes after TAVR, hospital length of stay and patient survival were analyzed according to NYHA classification. The median length of in-hospital stays following TAVR increased with higher NYHA functional classes (Figure 2). Specifically, patients classified as NYHA I/II had a median stay of 6.0 days (IQR 4.0-10.0), while NYHA III patients had a median inpatient stay of 7.0 days (IQR 5.0-10.0). NYHA IV patients remained in hospital for 10.0 days (IQR 6.0-13.0), indicating a statistically significant difference (p<0.001). 30-day mortality rates were higher with worsening NYHA class: 1.4% for NYHA I/II, 2.7% for NYHA III, and 8.3% for NYHA IV (p<0.001). This trend was consistent with the patterns observed for peri-interventional complications and length of in-hospital stay. Similarly, 1-year mortality rates showed a significant increase with NYHA classification, with 5.2% for NYHA I/II, 8.7% for NYHA III, and 19.2% for NYHA IV (p<0.001). These findings are summarized in Figure 3.

**Figure 2.**
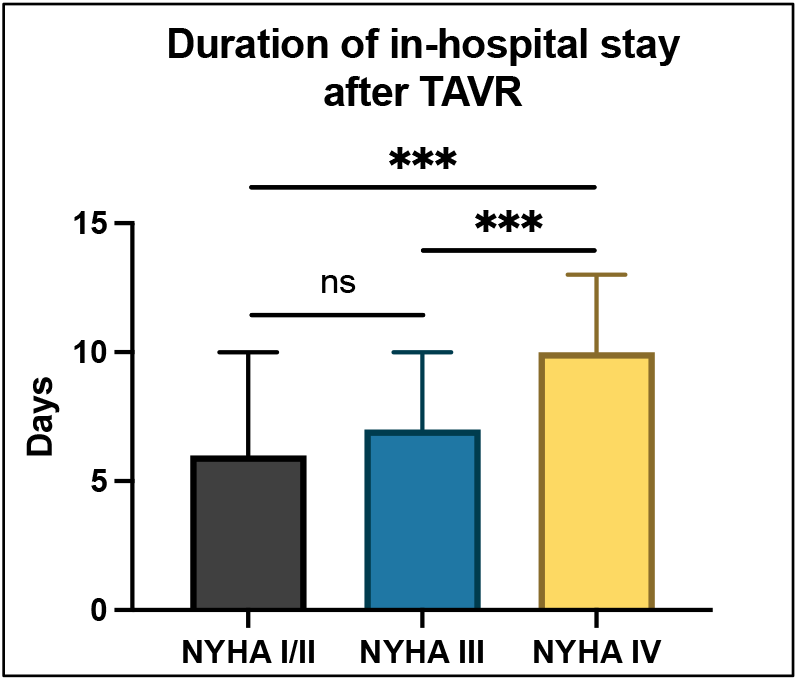
Duration of hospital stay following TAVR according to different NYHA classes. ns: not significant, ***p<0.001

**Figure 3.**
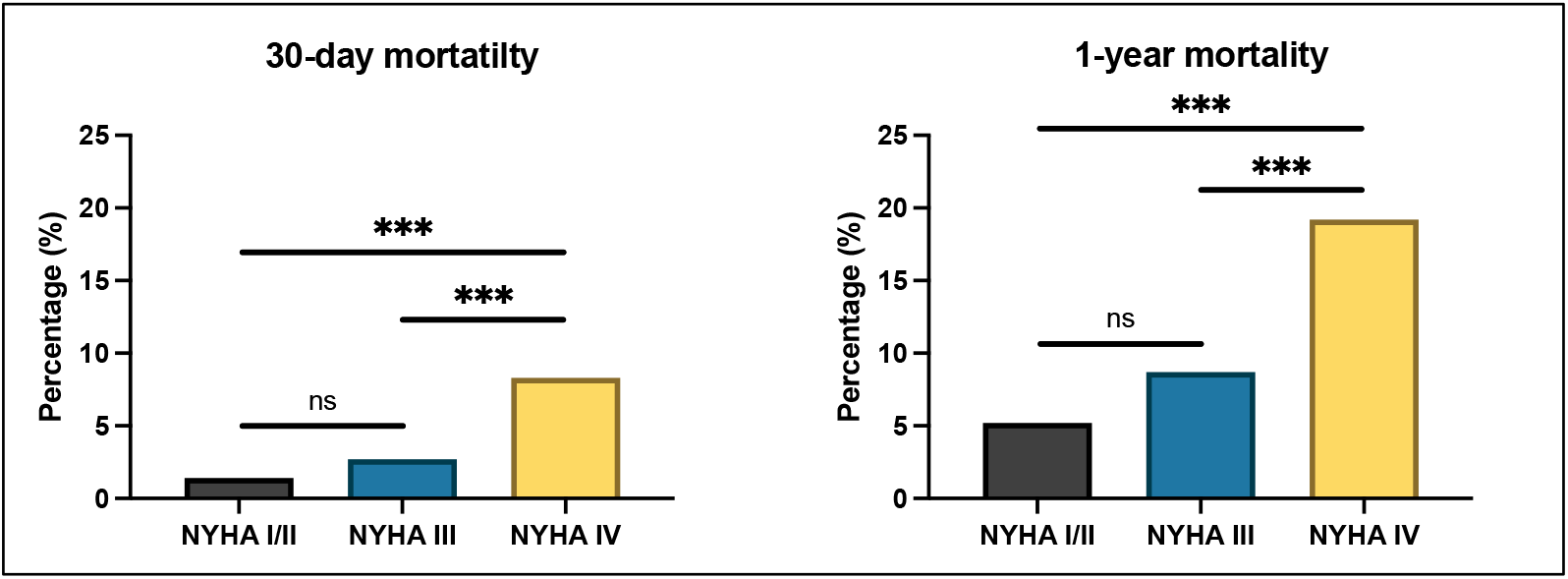
30-day and 1-year mortality following TAVR according to different NYHA classes. ns: not significant, ***p<0.001.

## Discussion

We here could demonstrate that pre-procedural NYHA classification is a strong predictor of peri-procedural complications, length of hospital stays, and both short- and long-term mortality after TAVR. These findings highlight the value of incorporating pre-procedural NYHA status into clinical decision-making. Furthermore, our data suggest that a comprehensive pre-interventional assessment, considering the patient’s NYHA classification, comorbidities, and overall condition, supports individualized treatment strategies, especially in high-risk TAVR patients.

A notable finding of our analysis is the high proportion of patients in advanced NYHA functional classes (III and IV) within the study cohort, representing a routine, unselected five-year series of TAVR cases treated at two high-volume German Heart Centers. While only 25.8% of patients were categorized in NYHA classes I and II, a striking 74.2% were classified in NYHA classes III and IV. This trend underscores the severity of the clinical condition in patients currently undergoing routine TAVR procedures. Given the ongoing demographic shift, characterized by an aging population and an increasing burden of comorbidities, the number of patients with advanced stages of HF requiring TAVR is expected to rise in the future. This observation not only highlights the growing complexity of TAVR indications but also underscores the need for more individualized management strategies to optimize outcomes in this high-risk cohort.^22^

As the rising number of elderly patients with advanced HF presents a particular challenge during clinical decision making, in our cohort, patients classified as NYHA IV were significantly older, with a mean age of 80.5 ± 6.3 years, compared to those in the NYHA I/II and III classifications (I/II: 79.1 ± 5.9 years, III: 79.7 ± 6.4 years; p=0.013). With advanced age, the prevalence of severe HF, right ventricular dysfunction, and frailty increases, potentially also reducing the ability to recover successfully after TAVR.^23,24^ Thus, a significant proportion of patients in our cohort presented with moderate to severe tricuspid regurgitation, itself associated with increased mortality (20.4% in NYHA III, 26.7% in NYHA IV vs. 17.1% in NYHA I/II; p=0.006). However, given the increasing life expectancy and the rising number of elderly patients, clinical decisions must place greater emphasis on the age-related comorbidity burden and focus on restoring the patient’s quality of life, particularly in the context of TAVR.^25^

Furthermore, our results demonstrate that patients in higher NYHA classes experienced significantly more peri-interventional complications. The need for prolonged vasopressor therapy (NYHA III: 5.5%, NYHA IV: 11.7% vs. NYHA I/II 1.6%), cardiopulmonary resuscitation (NYHA III: 2.2%, NYHA IV: 5.3% vs. NYHA I/II: 0.7%), and renal failure requiring RRT (NYHA III: 2.1%, NYHA IV: 6.8% vs. NYHA I/II: 0.2%), increased significantly with higher NYHA functional class (p**<**0.001). These complications, along with longer hospital stays, confirm that patients with advanced HF are at higher risk for immediate post-interventional challenges, presenting difficulties for healthcare professional teams and placing additional strain on healthcare resources and costs. By offering a detailed analysis of peri-interventional risk profiles across different NYHA classes, our study goes beyond existing literature in raising the question of whether patients in NYHA class IV should routinely undergo TAVR or, at best, receive targeted medical optimization prior to the procedure. While previous studies have established NYHA class as a prognostic marker, our data suggest that patients in NYHA class IV may have reached a clinical threshold beyond which TAVR may not automatically be beneficial, highlighting the need for particularly careful selection and more individualized pre-procedural preparation, especially in this vulnerable patient cohort.

Our data on long-term mortality finally confirm that the NYHA classification not only serves as an early risk parameter but also provides important prognostic value for long-term survival. 30-day mortality increased progressively with symptom severity, from 1.4% in NYHA I/II to 2.7% in NYHA III and 8.3% in NYHA IV (p<0.001). Similarly, the NYHA classification demonstrated strong prognostic value for long-term outcomes, with 1-year mortality rising from 5.2% in patients with NYHA class I/II to 8.7% in class III and 19.2% in class IV (p<0.001). These findings align with previous research, highlighting the impact of the overall symptom severity on post-TAVR outcomes.^8,26,27^ Moreover, this indicates that patients with advanced HF must be considered more carefully when selecting appropriate therapies to minimize the risk of poor outcomes.

Considering these findings, the role of the NYHA classification as a strong predictor in pre-procedural assessment for TAVR becomes increasingly important. A timely and integrated evaluation of the clinical status, comorbidities, and NYHA classification should serve as a cornerstone in the decision-making process when differentiating between TAVR and other treatment options. Particularly in older patients or those with advanced HF, the potential risks and expected post-TAVR quality of life should be factored into decision-making. A recent meta-analysis supports these findings and further demonstrates that urgent TAVR procedures in decompensated patients, compared to elective procedures, are associated with higher 30-day and in-hospital mortality, as well as increased vascular complications and acute renal failure.^28^ Future studies should investigate whether structured individualized pre-procedural stabilization strategies can improve outcomes in a growing high-risk population. Addressing both the clinical needs of an aging society and the economic challenges faced by healthcare systems, the focus should shift towards preemptive strategies, enabling therapeutic regimes before patients reach advanced stages of HF. In this context, efforts should be directed towards the identification of novel biomarkers and innovative targeted therapies, as well as enhanced and timely diagnostic approaches supported by artificial intelligence, such as advanced imaging techniques and personalized screening protocols, which could improve the early detection and monitoring of AS.^29,30^ Results from recent studies confirm that early TAVR intervention, even in still asymptomatic patients, may significantly improve survival and quality of life while preventing the progression to HF and decompensation.^31,32^

## Conclusions

Pre-procedural NYHA class is a strong and independent predictor of clinical outcomes in patients undergoing TAVR. Therefore, early intervention in cases of AS is essential to prevent clinical decompensation and to improve patient outcomes. However, in patients who present with higher NYHA classes, pre-procedural optimization strategies should be discussed. The incorporation of advanced diagnostic technologies and artificial intelligence offers promising opportunities for refined risk stratification and evidence-based decision-making. To minimize risk and increase procedural success, such individualized treatment strategies seem essential, particularly in an aging population with severe AS and concomitant HF. Given the increasing economic pressures on healthcare systems due to the growing number of elderly patients, optimizing patient management and procedural outcomes becomes even more critical to ensure the sustainability of healthcare resources.

## Limitations

This study has several limitations. The retrospective design and the limited number of contributing centers may restrict the generalizability of the findings and do not allow for causal inference. Pre-procedural NYHA class was assigned by healthcare professionals, making it susceptible to individual interpretation and potential variability. Objective measures of frailty or functional status were not included. Moreover, variations in procedural technique, device selection, and operator experience were not systematically captured. Important potential confounders, such as socioeconomic status, adherence to therapy, and access to follow-up care, were not assessed. Finally, clinical outcomes were not independently adjudicated, which may introduce reporting bias.

## Data Availability

Data will be available within the article or in an appropriate repository at time of publication.

## References

1. Vahanian A, Beyersdorf F, Praz F, et al. 2021 ESC/EACTS Guidelines for the management of valvular heart disease: Developed by the Task Force for the management of valvular heart disease of the European Society of Cardiology (ESC) and the European Association for Cardio-Thoracic Surgery (EACTS). European Heart Journal. 2022;43(7):561–632. doi:10.1093/eurheartj/ehab395

2. Leon MB, Smith CR, Mack MJ, et al. Transcatheter or Surgical Aortic-Valve Replacement in Intermediate-Risk Patients. N Engl J Med. 2016;374(17):1609–1620. doi:10.1056/NEJMoa1514616

3. Mack MJ, Leon MB, Thourani VH, et al. Transcatheter Aortic-Valve Replacement with a Balloon-Expandable Valve in Low-Risk Patients. N Engl J Med. 2019;380(18):1695–1705. doi:10.1056/NEJMoa1814052

4. Fischer-Rasokat U, Renker M, Charitos EI, et al. Cardiac decompensation of patients before transcatheter aortic valve implantation—clinical presentation, responsiveness to associated medication, and prognosis. Front Cardiovasc Med. 2023;10. doi:10.3389/fcvm.2023.1232054

5. Chen S, Redfors B, Crowley A, et al. Impact of recent heart failure hospitalization on clinical outcomes in patients with severe aortic stenosis undergoing transcatheter aortic valve replacement: an analysis from the PARTNER 2 trial and registries. Eur J Heart Fail. 2020;22(10):1866–1874. doi:10.1002/ejhf.1841

6. Strange JE, Nouhravesh N, Schou M, et al. High-risk admission prior to transcatheter aortic valve replacement and subsequent outcomes. Am Heart J. 2024;268:53–60. doi:10.1016/j.ahj.2023.11.003

7. McDonagh TA, Metra M, Adamo M, et al. 2021 ESC Guidelines for the diagnosis and treatment of acute and chronic heart failure: Developed by the Task Force for the diagnosis and treatment of acute and chronic heart failure of the European Society of Cardiology (ESC) With the special contribution of the Heart Failure Association (HFA) of the ESC. Rev Esp Cardiol (Engl Ed). 2022;75(6):523. doi:10.1016/j.rec.2022.05.005

8. Parikh PB, Mack M, Stone GW, et al. Transcatheter aortic valve replacement in heart failure. Eur J Heart Fail. 2024;26(2):460–470. doi:10.1002/ejhf.3151

9. Mengi S, Januzzi JL, Cavalcante JL, et al. Aortic Stenosis, Heart Failure, and Aortic Valve Replacement. JAMA Cardiol. 2024;9(12):1159–1168. doi:10.1001/jamacardio.2024.3486

10. Coisne A, Scotti A, Latib A, et al. Impact of Moderate Aortic Stenosis on Long-Term Clinical Outcomes: A Systematic Review and Meta-Analysis. JACC Cardiovasc Interv. 2022;15(16):1664–1674. doi:10.1016/j.jcin.2022.06.022

11. Généreux P, Schwartz A, Oldemeyer JB, et al. Transcatheter Aortic-Valve Replacement for Asymptomatic Severe Aortic Stenosis. N Engl J Med. Published online October 28, 2024. doi:10.1056/NEJMoa2405880

12. Mack MJ, Leon MB, Thourani VH, et al. Transcatheter Aortic-Valve Replacement in Low-Risk Patients at Five Years. N Engl J Med. 2023;389(21):1949–1960. doi:10.1056/NEJMoa2307447

13. Auffret V, Bakhti A, Leurent G, et al. Determinants and Impact of Heart Failure Readmission Following Transcatheter Aortic Valve Replacement. Circ Cardiovasc Interv. 2020;13(7):e008959. doi:10.1161/CIRCINTERVENTIONS.120.008959

14. Durand E, Doutriaux M, Bettinger N, et al. Incidence, Prognostic Impact, and Predictive Factors of Readmission for Heart Failure After Transcatheter Aortic Valve Replacement. JACC Cardiovasc Interv. 2017;10(23):2426–2436. doi:10.1016/j.jcin.2017.09.010

15. Leon MB, Smith CR, Mack M, et al. Transcatheter aortic-valve implantation for aortic stenosis in patients who cannot undergo surgery. N Engl J Med. 2010;363(17):1597–1607. doi:10.1056/NEJMoa1008232

16. Adams DH, Popma JJ, Reardon MJ, et al. Transcatheter aortic-valve replacement with a self-expanding prosthesis. N Engl J Med. 2014;370(19):1790–1798. doi:10.1056/NEJMoa1400590

17. Carroll JD, Mack MJ, Vemulapalli S, et al. STS-ACC TVT Registry of Transcatheter Aortic Valve Replacement. J Am Coll Cardiol. 2020;76(21):2492–2516. doi:10.1016/j.jacc.2020.09.595

18. Gilard M, Eltchaninoff H, Donzeau-Gouge P, et al. Late Outcomes of Transcatheter Aortic Valve Replacement in High-Risk Patients: The FRANCE-2 Registry. J Am Coll Cardiol. 2016;68(15):1637–1647. doi:10.1016/j.jacc.2016.07.747

19. Taniguchi T, Shirai S, Ando K, et al. Impact of New York Heart Association Functional Class on Outcomes After Transcatheter Aortic Valve Implantation. Cardiovasc Revasc Med. 2022;38:19–26. doi:10.1016/j.carrev.2021.07.022

20. Li S, Tang BY, Zhang B, et al. Analysis of risk factors and establishment of a risk prediction model for cardiothoracic surgical intensive care unit readmission after heart valve surgery in China: A single-center study. Heart Lung. 2019;48(1):61–68. doi:10.1016/j.hrtlng.2018.07.013

21. Magruder JT, Kashiouris M, Grimm JC, et al. A Predictive Model and Risk Score for Unplanned Cardiac Surgery Intensive Care Unit Readmissions. J Card Surg. 2015;30(9):685–690. doi:10.1111/jocs.12589

22. Bergmann T, Sengupta PP, Narula J. Is TAVR Ready for the Global Aging Population? Glob Heart. 2017;12(4):291–299. doi:10.1016/j.gheart.2017.02.002

23. Madanat L, Allam M, Khalili H, et al. Long-Term Survival and Quality of Life Following Transcatheter Aortic Valve Replacement in Nonagenarians. Am J Cardiol. 2024;213:140–145. doi:10.1016/j.amjcard.2023.12.031

24. Okoh AK, Chauhan D, Kang N, et al. The impact of frailty status on clinical and functional outcomes after transcatheter aortic valve replacement in nonagenarians with severe aortic stenosis. Catheter Cardiovasc Interv. 2017;90(6):1000–1006. doi:10.1002/ccd.27083

25. Galatas C, Afilalo J. Transcatheter aortic valve replacement over age 90: Risks vs benefits. Clin Cardiol. 2020;43(2):156–162. doi:10.1002/clc.23310

26. Adamo M, Fiorina C, Petronio AS, et al. Comparison of Early and Long-Term Outcomes After Transcatheter Aortic Valve Implantation in Patients with New York Heart Association Functional Class IV to those in Class III and Less. Am J Cardiol. 2018;122(10):1718–1726. doi:10.1016/j.amjcard.2018.08.006

27. Kim CA, Rasania SP, Afilalo J, Popma JJ, Lipsitz LA, Kim DH. Functional status and quality of life after transcatheter aortic valve replacement: a systematic review. Ann Intern Med. 2014;160(4):243–254. doi:10.7326/M13-1316

28. Apostolos A, Ktenopoulos N, Chlorogiannis DD, et al. Mortality Rates in Patients Undergoing Urgent Versus Elective Transcatheter Aortic Valve Replacement: A Meta-analysis. Angiology. Published online April 13, 2024:33197241245733. doi:10.1177/00033197241245733

29. Zhang Y, Wang M, Zhang E, Wu Y. Artificial Intelligence in the Screening, Diagnosis, and Management of Aortic Stenosis. Rev Cardiovasc Med. 2024;25(1):31. doi:10.31083/j.rcm2501031

30. Sun S, Yeh L, Imanzadeh A, Kooraki S, Kheradvar A, Bedayat A. The Current Landscape of Artificial Intelligence in Imaging for Transcatheter Aortic Valve Replacement. Curr Radiol Rep. 2024;12(11-12):113–120. doi:10.1007/s40134-024-00431-w

31. Beerkens FJ, Tang GHL, Kini AS, et al. Transcatheter Aortic Valve Replacement Beyond Severe Aortic Stenosis: JACC State-of-the-Art Review. J Am Coll Cardiol. 2025;85(9):944–964. doi:10.1016/j.jacc.2024.11.051

32. Généreux P, Schwartz A, Oldemeyer JB, et al. Transcatheter Aortic-Valve Replacement for Asymptomatic Severe Aortic Stenosis. N Engl J Med. 2025;392(3):217–227. doi:10.1056/NEJMoa2405880

